# Vascular burden and cognition: Mediating roles of neurodegeneration and amyloid-PET

**DOI:** 10.1101/2021.12.24.21267786

**Authors:** Julie Ottoy, Miracle Ozzoude, Katherine Zukotynski, Sabrina Adamo, Christopher Scott, Vincent Gaudet, Joel Ramirez, Walter Swardfager, Hugo Cogo-Moreira, Benjamin Lam, Aparna Bhan, Parisa Mojiri, Min Su Kang, Jennifer S. Rabin, Alex Kiss, Stephen Strother, Christian Bocti, Michael Borrie, Howard Chertkow, Richard Frayne, Robin Hsiung, Robert Laforce, Michael D. Noseworthy, Frank S. Prato, Demetrios J. Sahlas, Eric E. Smith, Phillip H. Kuo, Vesna Sossi, Alexander Thiel, Jean-Paul Soucy, Jean-Claude Tardif, Sandra E. Black, Maged Goubran, Medical Imaging Trials Network of Canada (MITNEC) and Alzheimer’s Disease Neuroimaging Initiative

**Author notes:** **Corresponding author:** Maged Goubran, Ph.D., 2075, 2075 Bayview Avenue, M6 West RM 176, Toronto, Canada, M4N 3M5. co-senior authors (Sandra E. Black, Maged Goubran). Data used in preparation of this article were obtained, in part, from the Alzheimer’s Disease Neuroimaging Initiative (ADNI) database (adni.loni.usc.edu). As such, the investigators within the ADNI contributed to the design and implementation of ADNI and/or provided data but did not participate in analysis or writing of this report. A complete listing of ADNI investigators can be found at: http://adni.loni.usc.edu/wp-content/uploads/how_to_apply/ADNI_Acknowledgement_List.pdf.

## Abstract

**INTRODUCTION:** It remains unclear to which extent vascular burden promotes neurodegeneration and cognitive dysfunction in a cohort spanning low-to-severe small vessel disease (SVD) and amyloid-beta pathology.

**METHODS:** In 120 subjects, we investigated 1) whether vascular burden, quantified as total or lobar white matter hyperintensity (WMH) volumes, is associated with different cognitive domains; and 2) whether the total WMH effect on cognition is mediated by amyloid (^18^F-AV45-PET), glucose metabolism (^18^F-FDG-PET), and/or cortical atrophy.

**RESULTS:** Increased total WMH volume was associated with poorer performance in all cognitive domains tested, with the strongest effects observed for semantic fluency. These relationships were mediated mainly through cortical atrophy, particularly in the temporal lobe, and to a lesser extent through amyloid and metabolism. WMH volumes differentially impacted cognition depending on lobar location and amyloid status.

**DISCUSSION:** Our study suggests mainly an amyloid-independent pathway in which vascular burden affects cognitive impairment through temporal lobe atrophy.

## 1. Introduction

White matter hyperintensities (WMH) detected on MRI are a common finding in stroke and dementia clinic patients [1,2]. Yet, the contributions of WMH to cognitive decline remain poorly understood. While Alzheimer’s disease (AD)-type pathologies such as amyloid-ß (Aß) plaques or tau neurofibrillary tangles are known to accelerate cognitive decline [3,4], the cognitive correlates of cerebral small vessel disease (SVD), specifically of WMH, have been both underestimated and understudied in AD [5].

SVD is a group of diseases that affects small arteries, venules, and capillaries of the brain [6]. MRI-based markers of SVD include WMH of presumed vascular origin, cerebral microbleeds, lacunae, and enlarged perivascular spaces. Amongst these markers, WMH is the most studied proxy of SVD. Notwithstanding that a low burden of WMH is frequently observed in healthy elderly [7,8], prior research indicated that WMH often co-exist with AD pathology [9]. In fact, more than 80% of AD dementia cases at autopsy may display cerebrovascular disease [10]. WMH are associated with an increased risk of stroke and (AD) dementia and faster clinical progression [11], highlighting that WMH should not just be interpreted as silent consequences of ageing [8].

Network studies have shown that focal WMH may initiate a cascade of events affecting remote brain areas often involved in AD [12,13]. In particular, WMH were found to be related to neurodegenerative processes (hypometabolism/atrophy) in AD-vulnerable regions [14–20]. Specifically, these neurodegenerative processes were demonstrated to *mediate* the relationship between SVD burden and cognitive impairment [15,18,21]. Longitudinal studies, in turn, supported the potential causal relationship between SVD and cognitive impairment by showing that elevated WMH burden at baseline worsened neurodegeneration and cognition over time in AD cohorts [22,23]. Similarly, studies have highlighted that baseline WMH predicted higher Aß deposition over time [24–26], and even suggested that vascular abnormality precedes Aß deposition in the cascade of AD pathophysiology [27].

However, hitherto, the majority of large-cohort studies in dementia that investigated the effects of WMH on neurodegeneration and cognition excluded subjects with considerable WMH burden, such as the Alzheimer’s Disease Neuroimaging Initiative (ADNI). Others focused on cognitively normal elderly, with few enriched for high WMH burden [15]. Therefore, our knowledge is still limited regarding the effects of vascular pathology in patients of more extreme endophenotypes with evidence of significant WMH burden in addition to AD pathology [5]. Furthermore, to our knowledge, no studies have yet comprehensively studied the potential mediating roles of Aß, glucose metabolism, and atrophy in the vascular contributions to cognitive impairment [26].

In this study, we investigated a unique cohort of clinically normal elderly and “real-world” patients capturing the spectrum of low to extensive WM disease and Aß pathology. The objectives were two-fold: 1) investigate whether vascular burden, quantified as total or lobar WMH volume, is associated with cognition; 2) investigate the potential mediating roles of Aß, glucose metabolism, and/or cortical atrophy in the WMH-cognition relationship.

## 2. Methods

### 2.1 Participants

The study included 120 subjects in total. Sixty subjects were recruited in a multicenter prospective observational study through seven participating sites as part of the C6 project in the Medical Imaging Trials Network of Canada (MITNEC-C6). They were enrolled from stroke-prevention clinics (i.e., transient ischemic attacks or minor subcortical lacunar infarcts) and dementia clinics and presented with moderate-to-severe SVD as quantified by Fazekas-score>2 and high (>10cm^3^[28]) confluent periventricular WMH volumes [median(IQR): 30.51(22.14)cm^3^]. Detailed selection criteria are described in **Supplementary Table 1** and in Zukotynski et al.[29]. In addition, the study included sixty cognitively normal and mild cognitive impairment (MCI) subjects from the baseline ADNI-2 database with low-to-moderate WMH [median(IQR): 5.82(9.29)cm^3^]. Both cohorts were well matched for vascular risk factors including hypertension, pulse pressure, BMI, sex, and smoking status. Detailed demographics are reported in **Supplementary Table 2**. The institutional review boards at all participating institutions approved this study and all participants provided written informed consent.

### 2.2 Assessments

A battery of cognitive tests was administered including: processing speed (Trail Making Test part-A, n=120); executive function (Trail Making Test part-B, n=119); semantic fluency (animal naming, n=120); and language (Boston Naming Test or BNT, n=120). Exploratory analyses included global cognition (Montreal Cognitive Assessment or MoCA, n=120) and global function (Functional Assessment Questionnaire or FAQ, n=110). Individual assessments are described in **Supplementary Table 3**.

### 2.3 Neuroimaging

Each subject underwent a 3T MRI scan, including 3D T1 and fluid-attenuated inversion recovery (FLAIR) structural sequences. Acquisition parameters are described in **Supplementary Table 4** and followed a common imaging protocol [30]. Each subject also underwent a ^18^F-florbetapir (^18^F-AV45) PET scan to quantify brain Aß (n=120) and a ^18^F-fluorodeoxyglucose (^18^F-FDG) PET scan (n=117) to quantify brain glucose metabolism. Consistency of PET data between participating sites was maintained by use of the main ADNI-2 PET protocol [see http://adni.loni.usc.edu/wp-content/uploads/2008/07/adni2-proceduresmanual.pdf]. All imaging data was transferred to a central site for final quality check.

ADNI was launched in 2003 as a public-private partnership. The primary goal of ADNI was to test whether MRI, PET, other biological markers, and clinical data can be combined to measure the progression of MCI/AD [see www.adni-info.org].

### 2.4 MRI processing

Atrophy was quantified using cortical thickness, extracted based on T1-weighted images using FreeSurfer v6.0. We employed a modified FreeSurfer workflow for patients with WMH burden [31]. An ‘AD-signature’ thickness meta-ROI was employed based on surface-weighted thickness averages of the entorhinal, fusiform, parahippocampal, mid/inferior temporal, and inferior parietal gyri [32].

WMH were delineated based on our automated segmentation tool ‘HyperMapper’ that uses T1, FLAIR, and Bayesian 3D Convolutional Neuronal Networks (CNN) [33]. Subcortical lacunar infarcts were masked before WMH segmentation [34]. Intracranial volume (ICV) was extracted using our CNN-based ‘iCVMapper’ tool, which was shown to be more robust compared to other state-of-the-art skull-stripping methods [35]. Finally, vascular burden was defined as WMH volume divided by ICV and log-transformed.

Lobar WMH volumes were determined by intersecting the total WMH mask and each individual lobar WM mask, as delineated in native T1 space based on the Desikan-Killiany-Tourville (DKT) atlas of FreeSurfer. Lobar WM masks included the frontal, parietal, temporal, occipital, insula, and cingulate lobes. **Figure 1** shows examples of our structural MRI scans, WMH and lobar delineations, as well as a heatmap of WMH volumes across the MITNEC-C6 cohort.

**Fig 1.**
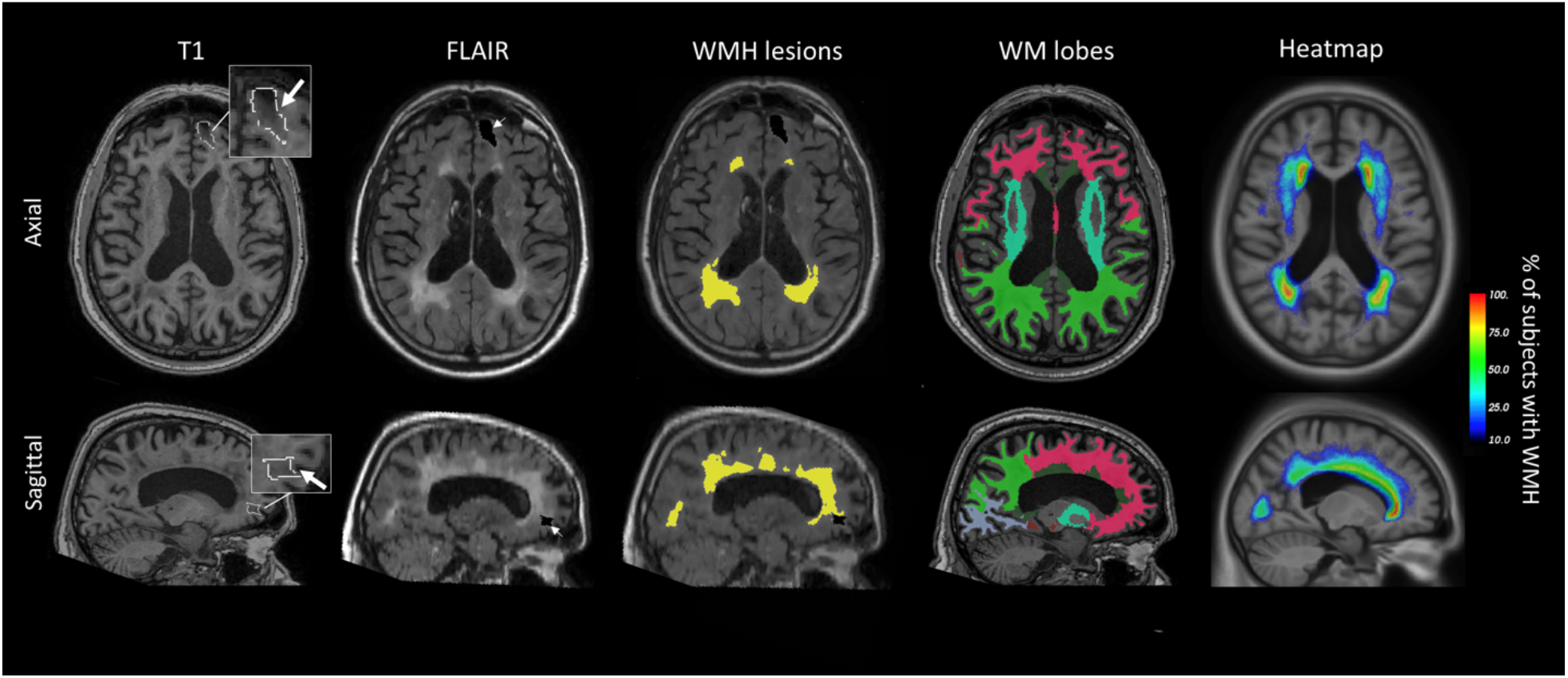
Volumetric delineations. Example of brain segmentation for one subject (age range 60-70 years, male) and WMH heatmap across subjects (outer right column). Left to right: T1 with delineation of stroke region; FLAIR with the stroke region masked out; FLAIR with WMH delineations in yellow color; WM lobe segmentation [pink: frontal, brown: temporal, light green: parietal, purple: occipital; cyan: insula; dark green: cingulate]; WMH volume distribution ‘heatmap’ across the MITNEC-C6 subjects (% of subjects with WMH per voxel) overlayed on the ADNI template.

### 2.5 PET Image processing

PET images were processed using PetSurfer v6.0. This included motion-correction of the individual PET frames to the first frame and averaging to obtain one static frame, co-registration to T1-weighted image, smoothing to 8mm full-width-at-half-maximum to reach a common resolution across sites [36], and generating standardized uptake value ratios (SUVR) maps. The ^18^F-AV45 and ^18^F-FDG SUVR maps were referenced to the cerebellum and the pons, respectively [37]. Partial volume correction (PVC) was applied using the geometric-transfer-matrix method with a point-spread-function of 8mm. Regional values were extracted based on the DKT atlas in native T1 space. AD-signature meta-ROIs were created for ^18^F-AV45 and ^18^F-FDG SUVR based on Jack’s mask (volume-weighted average of the frontal, parietal, temporal, and cingulate regions) and the temporo-parietal lobe, respectively [37–39]. Based on our Aß-PET pipeline, 22% and 48% of ADNI and MITNEC-C6 subjects, respectively, were considered Aß-positive using a quantitative ^18^F-AV45 SUVR cut-off of 1.1. This cut-off was derived from Gaussian mixture modelling with two components using non-PVC SUVR in the AD-signature meta-ROI. Similar results were found when using PVC data.

### 2.6 Statistical analyses

Regional statistics were performed in SPSS v24 (SPSS Inc., Chicago, IL) and vertex-wise statistics were performed in FreeSurfer v6.0. All metrics were z-scored to allow for direct comparison between models across predictors and outcome measures. Values were reported as mean ± standard error (SE), unless otherwise stated.

#### 2.6.1 Group differences

A two-tailed t-test for continuous variables and chi-square for categorical variables were used to detect significant group differences in the demographics. A two-way ANOVA (lobe*group), corrected for age, sex, education, and Sidak’s multiple comparisons test, was used to detect group differences in lobar WMH volumes.

#### 2.6.2 Regressions

Linear regressions were employed to assess the associations between total/lobar WMH volume (independent variable) and each of the cognitive scores (dependent variable), adjusted for age, sex, and education. Bias-corrected bootstrapping with 5,000 replications was applied to account for multiple comparisons and heteroscedasticity. Bias-corrected bootstrapping does not make any assumptions about normality in the sampling distribution and better controls type I errors.

#### 2.6.3 Mediation analyses

For mediation analyses, the PROCESS macro v3.5 in SPSS was applied. Bias-corrected bootstrapping with 5,000 replications was performed for estimation of (in)direct and total effects. We hypothesized that a serial mediation runs from ‘total WMH →Aß [24,25] → atrophy [22,23] → cognition’. Importantly, this mediation model allows not only to investigate indirect effects of WMH volumes on cognition through the hypothesized serial path, but also through the predictor and each of the mediators separately while adjusting for the remaining variables in the model. Due to the previously reported association of ^18^F-FDG-PET with semantic fluency and executive function [40], glucose metabolism was tested as an additional mediator within their path analyses: total WMH → Aß → metabolism [20,41] → atrophy [42] → fluency or executive function; the cited references indicate prior research establishing this sequence of biomarkers becoming abnormal in AD. Imaging mediators were evaluated within AD-signature regions (see section 2.5). Age, education, and sex were used as covariates regressed on the mediators and outcome simultaneously.

To test regional specificity of the mediation analyses, we performed additional path analysis whereby AD-signature meta-ROIs of the imaging markers were substituted by a more focal region. This region was selected through two separate whole-brain vertex-wise linear regressions of total WMH volume with (i) atrophy [cortical thickness] and (ii) Aß SUVR, to identify one ‘WMH-signature’ region in relation to both atrophy and Aß. These vertex-wise regressions were adjusted for age, sex, education, and (i) additionally for global Aß SUVR. Vertex-wise multiple comparisons correction was based on Monte-Carlo simulations with 5,000 iterations, which implemented a two-tailed cluster-forming *P*-value of 0.01.

## 3. Results

### 3.1 Associations between WMH and cognition

**Table 1** represents the demographics within the low and high WMH groups. Greater total WMH volume was associated with poorer performance on the following assessments across all subjects in rank order of effect size: semantic fluency (β=-0.35±0.09, *P*=0.0004), executive function (β=+0.34±0.09, *P*=0.0004), global function (β=+0.30±0.10, *P*=0.005), processing speed (β=+0.22±0.08, *P*=0.013), and global cognition (β=-0.17±0.08, *P*=0.040) (**Figure 2**A). No significant associations were found with the language domain as assessed by BNT (*P*=0.13).

**Table 1.** Demographics. Demographics within the low and high WMH groups, determined based on standard binning. All values are indicated as mean ± standard deviation except for sex and Aß-positivity (^*^P<0.01, ^**^P<0.0001). Abbreviations: MMSE, mini-mental state examination; MoCA, Montreal cognitive assessment; SUVR, standardized uptake value ratio; WMH, white matter hyperintensity volumes.

| **Variables** | **Low WMH group**<br>**(n = 60)** | **High WMH group**<br>**(n = 60)** | *Test-statistic* |
| --- | --- | --- | --- |
| **Age (years)** | 72.85 ± 5.96 | 78.00 ± 7.01 | t = -4.3 ** |
| **Sex male, n (%)** | 29 (48%) | 30 (50%) | $\chi^2$ = 0.03 |
| **Education (years)** | 15.67 ± 2.87 | 14.72 ± 2.76 | t = 1.9 |
| **Composite [<sup>18</sup>F]AV45 SUVR\*** | 0.74 ± 0.32 | 1.08 ± 0.46* | t = -4.6 ** |
| **Composite [<sup>18</sup>F]FDG SUVR\*** | 2.32 ± 0.27 | 2.24 ± 0.36 | t = 1.3 |
| **Cortical thickness (global, mm)** | 2.34 ± 0.09 | 2.25 ± 0.09 | t = 5.3 ** |
| **Total WMH (cm<sup>3</sup>)** | 7.54 ± 5.14 | 37.22 ± 18.09 | t = -12.2 ** |
| **MMSE** | 28.75 ± 1.68 | 27.22 ± 2.39 | t = 4.1 ** |
| **MoCA** | 25.53 ± 3.12 | 22.5 ± 4.04 | t = 4.6 ** |

\* SUVR values were corrected for partial volume effects. Composite [<sup>18</sup>F]AV45 and [<sup>18</sup>F]FDG SUVR were based on volume-weighted averages of AD-signature regions [37,39].

**Fig 2.**
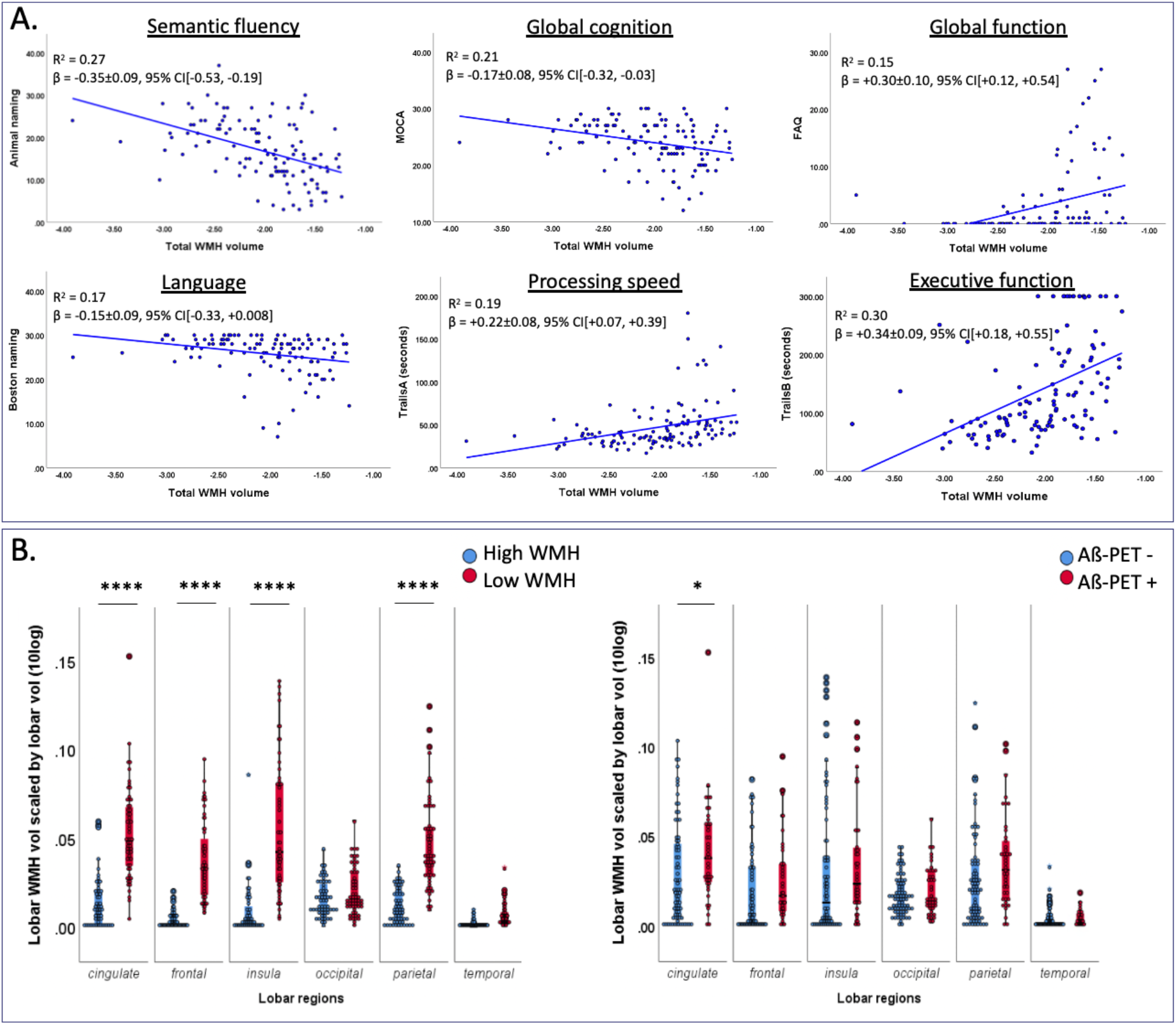
Total and lobar WMH volumes. A) Linear regressions between cognitive test scores and total WMH volumes across all subjects. Confidence intervals (95% CI) are bootstrapped with 5,000 replications and adjusted for age, sex, and education. B) Lobar WMH volumes in the high vs low total WMH group (left) and in the Aß-positive vs negative group (right). P-values were based on a two-way ANOVA that investigated whether lobar location and group influenced WMH burden [54]. Values were adjusted for multiple comparisons and age, sex, and education (^*^P<0.05, ^**^P<0.01, ^***^P<0.001, ^****^P<0.0001). Abbreviations: FAQ, functional assessment questionnaire; MoCA, Montreal cognitive assessment; WMH, white matter hyperintensity volumes.

Volumes of lobar WMH were significantly higher in the high WMH group in the frontal, parietal, cingulate, and insula lobes (**Figure 2**B). Cognitive correlates of *frontal* WMH volumes were significant only in Aß-negative subjects, whereas cognitive correlates of *temporal* and *parietal/cingulate* WMH volumes were more prominent in the Aß-positive subjects (**Supplementary Table 5**). Only *cingulate* WMH volumes were significantly increased in the Aß-positive compared to the Aß-negative group (**Figure 2**B).

### 3.2 Aß, metabolism, and cortical thickness mediate the effects of WMH on cognition

#### 3.2.1 AD-signature meta-ROIs

In the path analysis for semantic fluency, we found significant indirect effects of total WMH volume through (i) ‘WMH→atrophy→fluency’, (ii) ‘WMH→Aß→atrophy→fluency’, and (iii) ‘WMH→Aß→hypometabolism→fluency’ (total of 68% mediation, β=-0.23±0.06, 95% CI[-0.36,-0.10]) (**Figure 3**A). Path (i) via atrophy explained most of the indirect effect (β=-0.20±0.05, 95% CI[-0.31,-0.11]). The direct effect of WMH volume on semantic fluency (i.e., after controlling for the mediators and covariates) became non-significant.

**Fig 3.**
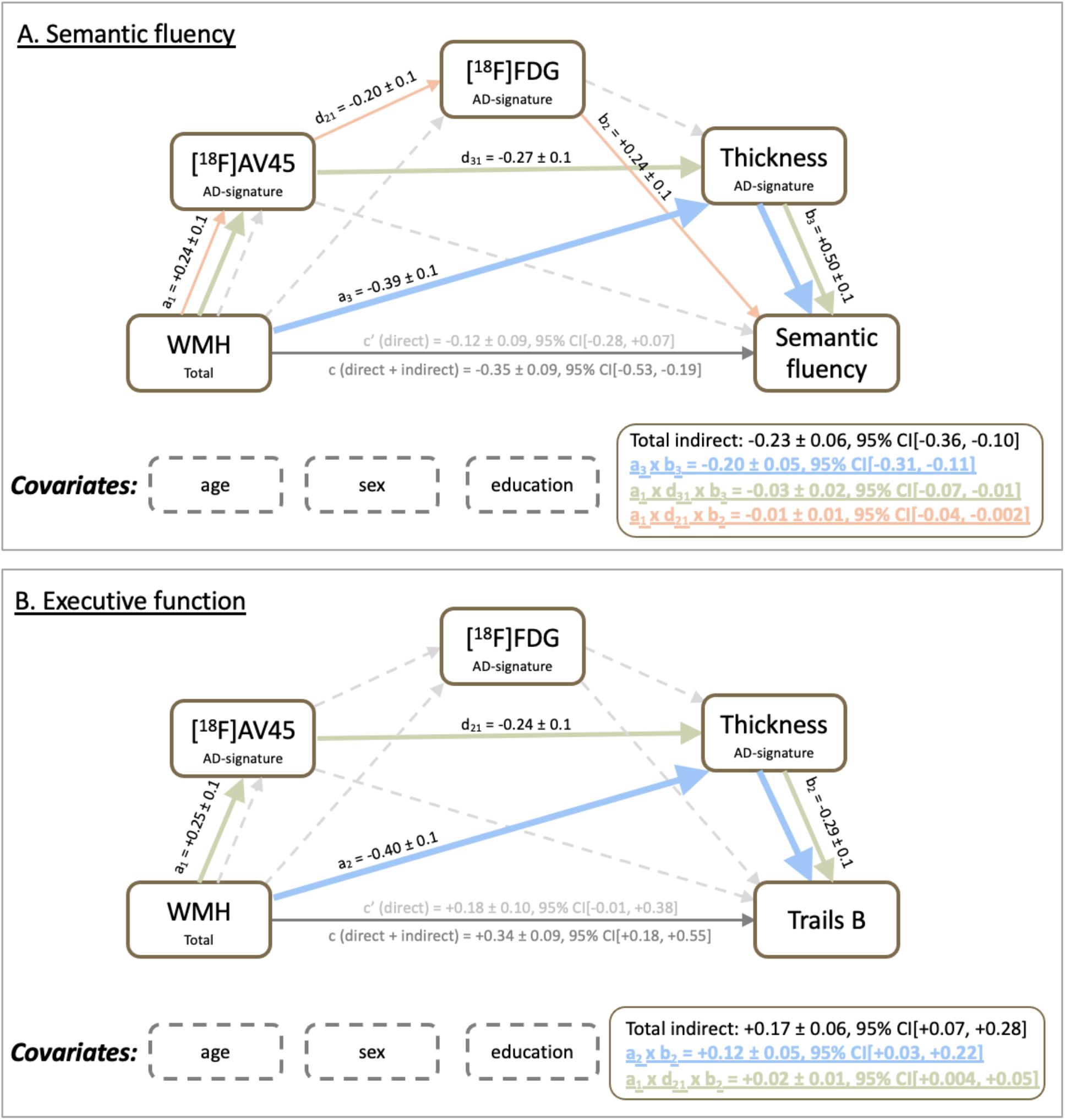
Mediation analyses of Aß, metabolism and atrophy on the WMH-cognition relationship. Mediation models showing significant paths mediating the association of total WMH volumes with semantic fluency (Panel A) and executive function (Panel B). Thick lines are part of a significant pathway, whereas dashed lines represent non-significant pathways. All mediators used an AD-signature meta-ROI [32,37,39]. Values are indicated as mean ± SE and 95% CI are bootstrapped with 5,000 replications. Path c represents the total (direct + indirect) effect adjusted only for covariates, whereas c’ represents the direct effect adjusted for covariates and indirect effects.

A similar path result was found for executive function (i.e., with cortical atrophy explaining most of the indirect effect), though ^18^F-FDG PET was not a significant mediator. The total indirect effects via atrophy and Aß corresponded to 50% (β=+0.17±0.06, 95% CI[+0.07, +0.28]) (**Figure 3**B). Exploratory path analyses of global function and global cognition through atrophy and Aß are reported in **Supplementary Figure 1**. The direct effect of WMH volume on these cognitive metrics similarly became non-significant due to mediation. No significant mediating effects were detected for processing speed (β=+0.10±0.06, 95% CI[-0.006,+0.23]).

#### 3.2.2 Temporal meta-ROI

Vertex-wise regression analyses showed that total WMH volume was positively associated with Aß load and inversely associated with atrophy, particularly in lateral temporal regions (**Figure 4**A). As such, we performed further path analysis on cognition using a temporal meta-ROI for all mediators. The temporal meta-ROI consisted of the inferior, superior, and middle temporal lobe and the temporal pole. When using a temporal meta-ROI for each of the mediators, we found higher effect sizes of the mediating paths compared to the model based on AD-signature meta-ROIs (**Figure 4**B). The total indirect effects via atrophy, Aß, and metabolism corresponded to 74% for semantic fluency (β = -0.26±0.07, 95% CI[-0.39,-0.13]), while the total indirect effects via atrophy and Aß corresponded to 62% for executive function (β=+0.21±0.06, 95% CI[+0.10,+0.33]) (**Figure 4**B). Exploratory path analyses of global function, global cognition, and processing speed are reported in **Supplementary Figure** 2. Similar to the AD-signature regions results, the indirect path ‘WMH→atrophy→cognition’ explained most of the indirect effect.

**Fig 4.**
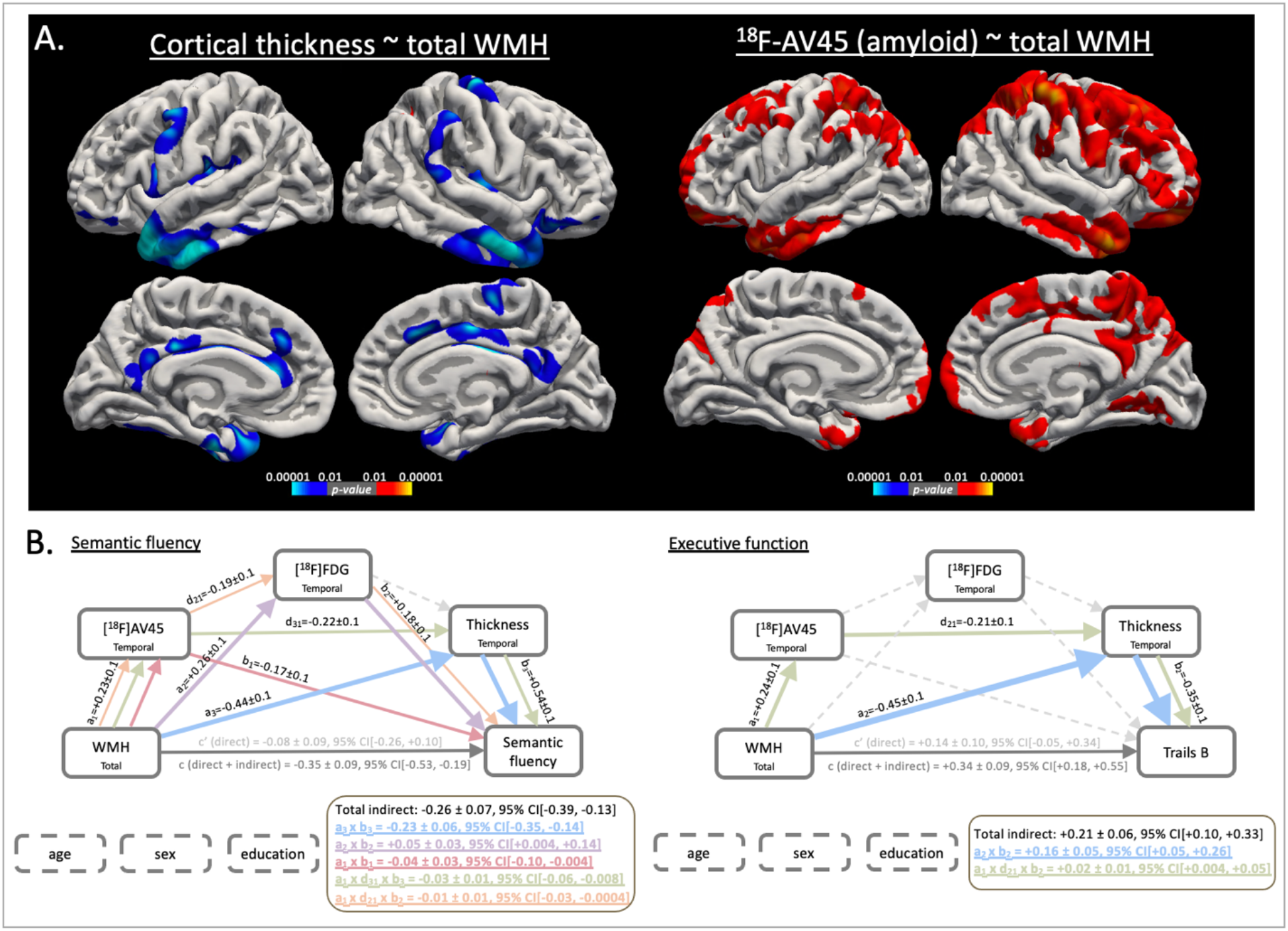
Temporal lobe-focused mediation analyses based on vertex-wise associations of WMH burden with atrophy and Aß. (Panel A) Vertex-wise regression analysis of total WMH volumes with cortical thickness (left) and ^18^F-AV45 Aß PET (right) across all subjects. Blue and red represent negative and positive associations respectively. Both modalities showed the strongest association with total WMH volumes in the temporal lobe, which was then used as a meta-ROI in subsequent mediation analyses, see Panel B. Results are displayed at P<0.01 after cluster-wise correction for multiple comparisons. (Panel B) Mediation models showing which paths are significantly mediating the association of total WMH volumes with semantic fluency (left) and executive function (right). Thick lines are part of a significant pathway, whereas dashed lines represent non-significant pathways. All mediators used a temporal meta-ROI. Values are indicated as mean ± SE and 95% CI are bootstrapped with 5,000 replications. Path c represents the total (direct + indirect) effect adjusted only for covariates, whereas c’ represents the direct effect adjusted for covariates and indirect effects.

### 3.3 Post-hoc mediation model

A post-hoc mediation model was investigated using Aß as the predictor and WMH as the mediator: ‘Aß→WMH→atrophy→cognition’. Rather than investigating the causal relationships between Aß and WMH, this model was applied to support our previous findings that the indirect effects of Aß and WMH through each other on atrophy and cognition have lower effect size compared to the effect of Aß or WMH alone.

Similar significant paths (**Supplementary Figure 3**) were found as described above for each of the cognitive domains (see section 3.2). Importantly, the indirect path ‘Aß→atrophy→cognition’ had larger effect size than its effect through WMH (‘Aß→WMH→atrophy→cognition’). This paralleled the results described above (see section 3.2), where the indirect path ‘WMH→atrophy→cognition’ had larger effect size than its effect through Aß (‘WMH→Aß→atrophy→cognition’). Taken together, this may suggest that WMH and Aß exert more additive and less synergistic effects on atrophy (i.e., potentially driving further atrophy through different underlying mechanisms).

Furthermore, the Aß-independent effect of WMH on atrophy was greater than the WMH-independent effect of Aß on atrophy in both the initial and post-hoc models. Taken together, this may suggest that the effect of WMH on atrophy exceeds the effect of Aß on atrophy.

## 4. Discussion

This study investigated the relationship between WMH burden and standardized neuropsychological assessments in a unique cohort of clinically normal elderly and “real-world patients” capturing the spectrum of low to extensive WM disease and Aß pathology. We found that WMH volumes were associated with poorer semantic fluency, executive function, and global function. These relationships were mostly due to the association of WMH volumes with cortical atrophy, particularly in the temporal lobe, and to a lesser extent due to the association of WMH volumes with Aß or glucose metabolism.

The contribution of WMH to cognitive deficits has been thus far poorly understood. Previous literature showed direct, indirect, and no effects of WMH on cognitive decline in various populations [26,43]. Insofar, most large AD/dementia cohort studies excluded patients with considerate amount of vascular co-pathology as mixed disease. As such, potential contributions of WMH to cognitive impairment are often not well addressed in these “clean” cases of probable AD [7]. While a considerate portion of both cognitively normal and AD dementia cases display WMH [8,10,28], the prevalence of WMH is substantially higher in stroke and dementia clinic patients [1,2]. This group of patients thus provides an enriched source for recruitment of more extreme vascular endophenotypes. In our WMH-enriched cohort (totaling n=120), we found an association between WMH volumes and cognitive impairment but not after controlling for atrophy, particularly of the temporal lobe. This suggests an indirect (‘mediation’) effect of WMH on cognition through neurodegenerative processes. Similarly, studies have shown that WMH contribute to atrophy beyond age-related effects, particularly in the frontal and temporal lobes [14,16], and that the effect of WMH volumes on cognition is mediated by atrophy [15,18,21]. Our findings may imply that imaging-visible WM lesions exert secondary widespread and neurodegenerative effects on the grey matter by damaging specific WM tracts that subserve these cortical regions. The (anterior) temporal lobe may be especially vulnerable to atrophy in WMH-enriched dementia cohorts because of its susceptibility to both AD-and vascular-related damage [44] and the potential vulnerability of connecting tracts to WMH crossing, such as the uncinate fasciculus [45].

Unlike previous studies, we employed PET-based biomarkers of Aß and glucose metabolism in addition to atrophy as potential mediators in the model. This allowed us to investigate whether effects of WMH volumes on cognition were also indirectly promoted by Aß or glucose metabolism. Indeed, additional indirect paths via Aß followed by neurodegeneration (metabolism or atrophy) mediated the WMH-cognition relationship. These indirect paths through Aß, however, had lower effect size, suggesting that WMH and AD pathophysiology exert mostly additive effects on cognitive impairment. The more prominent additive effects were also underscored by our alternative path model, where Aß affected cognition mostly through cortical atrophy rather than through WMH. Our findings are in line with prior studies reporting additive effects of WMH and AD pathology on cognitive decline at baseline and longitudinally [46,47], as well as a lower degree of AD pathology in the presence of vascular pathology for the same level of cognitive impairment [48] and a substantial increase in dementia risk post-stroke [49]. Importantly, apart from additive effects, we also show that the indirect path through Aß (i.e., ‘WMH→Aß→(..)→cognition’) was significant. As such, subjects with elevated WMH burden may represent an at-risk group not only for increased neurodegeneration but *also* for Aß deposition [24]. Thus, subjects with vascular co-pathology may represent an important target group for dementia prevention when vascular risk factors can be treated alongside AD hallmarks.

In relation to cognition, we showed that higher volumes of WMH correlated with slower processing (psychomotor) speed and impaired executive function [11]. While processing speed was directly impacted by WMH burden, we found that executive function was impacted only *indirectly* mainly through cortical atrophy. Another strong correlate of WMH in our study was semantic fluency, one of the earliest domains to become impaired in AD [40] and sensitive to discriminate AD/vascular dementia from normal aging [50]. Interestingly, we found that neurodegeneration of the temporal/AD-signature region was a stronger mediator in the WMH-fluency relationship than in the WMH-executive relationship. Indeed, semantic fluency is sustained primarily by the left temporal lobe, with language processing being a critical component for this task [45], while executive function is thought to be predominantly frontal-mediated (not being a region-of-focus in the current study).

While numerous studies reported brain-behaviour relationships using total WMH volumes, only a few focused on the role of regional (lobar) WMH. Our exploratory analyses support the hypothesis that regional WMH burden may differentially affect cognition [43], and highlighted that this relationship may depend on Aß status. Specifically, *frontal* WMH volumes were associated with poorer processing speed, semantic fluency, and executive function, particularly in the normal (Aß-negative) subjects. Previous research likewise found an association of *frontal* WMH volumes with the so-called dysexecutive syndrome in normal elderly subjects [43]. Conversely, *cingulate* and *temporal* WMH volumes played a more prominent role in the Aß-positive subjects in our study. Indeed, emerging evidence has shown vulnerability of the cingulate and temporal areas in the relationship between AD and vascular pathology. For example, Van Westen and colleagues [9] reported an association between *temporal* WMH and elevated Aß using both fluid and PET-based biomarkers in AD dementia. Second, McAleese et al. [51] reported that parietal/cingulate WMH are related to AD rather than to SVD. While the latter authors concluded that WMH is primarily a consequence of Wallerian degeneration triggered by cortical AD-type pathology itself, other studies have supported the idea that SVD exacerbates AD-related pathology by inducing neuroinflammatory responses and by reducing the clearance of toxic proteins (including Aß) from the brain [26]. In fact, our group previously showed that collagenosis of the large deep medullary veins is strongly associated with periventricular WMH burden in patients with and without AD [52]. Longitudinal studies may help to establish potential causal relationships between WMH and AD pathology.

The present study had several limitations. First, our study had relatively small sample size (n=120), due in part to recruitment of a unique cohort of “real-world” patients who demonstrated moderate-to-severe WM disease. Therefore, we limited the number of mediation analyses by investigating 1) all subjects combined and 2) using total (rather than lobar) WMH volumes. Second, we did not include a marker of hyperphosphorylated tau, which is expected to be associated with atrophy. The relationship between tau pathology and vascular burden remains debated in literature, with most of the in-vivo biomarker studies not reporting an association [53]. Furthermore, we found a sustained association between WMH volumes and cognitive deficits in the Aß-negative subjects who are unlikely to have substantial AD-related tau pathology (see supplementary Table 2), indicating that the impact of WMH on cognition may not strongly depend on AD-related tauopathy.

The strengths of this study lie in the use of a broad dynamic range of WMH burden and cognitive scores by the inclusion of mixed SVD and AD pathologies, the addition of PET-based biomarkers of Aß and glucose metabolism, as well as the use of optimized segmentation tools to determine WMH volumes and cortical thickness. In conclusion, our study suggests that increased WMH volumes affect cognitive impairment mainly through cortical atrophy and to a lesser extent through alterations in Aß deposition and glucose metabolism.

## Data Availability

All data associated with this study are available in the main text or the supplementary materials. All the imaging data can be shared upon request with a proposal and under a data transfer agreement.

## Acknowledgements

We would like to express our deepest gratitude towards all the participants and caregivers for their support and participation in this study. We are grateful for the support of the Medical Imaging Trial Network of Canada (MITNEC) Grant #NCT02330510, and to Eli Lilly & Company for providing the 18F-florbetapir ligand. In addition, part of the data collection and sharing for this project was funded by ADNI (National Institutes of Health Grant U01 AG024904) and DOD ADNI (Department of Defense award number W81XWH-12-2-0012). ADNI is funded by the National Institute on Aging, the National Institute of Biomedical Imaging and Bioengineering, and through generous contributions from the following: AbbVie, Alzheimer’s Association; Alzheimer’s Drug Discovery Foundation; Araclon Biotech; BioClinica, Inc.; Biogen; Bristol-Myers Squibb Company; CereSpir, Inc.; Cogstate; Eisai Inc.; Elan Pharmaceuticals, Inc.; Eli Lilly and Company; EuroImmun; F. Hoffmann-La Roche Ltd and its affiliated company Genentech, Inc.; Fujirebio; GE Healthcare; IXICO Ltd.; Janssen Alzheimer Immunotherapy Research & Development, LLC.; Johnson & Johnson Pharmaceutical Research & Development LLC.; Lumosity; Lundbeck; Merck & Co., Inc.; Meso Scale Diagnostics, LLC.; NeuroRx Research; Neurotrack Technologies; Novartis Pharmaceuticals Corporation; Pfizer Inc.; Piramal Imaging; Servier; Takeda Pharmaceutical Company; and Transition Therapeutics. The Canadian Institutes of Health Research provided some funding to support ADNI clinical sites in Canada. Private sector contributions are facilitated by the Foundation for the National Institutes of Health (www.fnih.org). The grantee organization is the Northern California Institute for Research and Education, and the study is coordinated by the Alzheimer’s Therapeutic Research Institute at the University of Southern California. ADNI data are disseminated by the Laboratory for Neuro Imaging at the University of Southern California.

## Author contributions

JO, SEB and MG designed the study and experiments. JO and MG wrote the manuscript. JO analyzed and interpreted the data with input from MG and SEB. MO, KZ, HCM, WS, JSR, revised the manuscript, participated in experiment design, data analysis and interpretation. MSK, PHK, SA, VG, CS, JR, AP, PM, and BL contributed to data generation and manuscript revision. AK, SS, CB, MB, HC, RF, RH, RJL, MDN, FSP, DJS, EES, VS, AT, JPS, and JCT participated in study concept and design as site-leaders and revised the manuscript. MG and SEB supervised all aspects of this work.

## Supplementary Materials

1. Fig S1. Mediation analyses of WMH on global function and global cognition, using AD-signature meta-ROIs.
2. Fig S2. Mediation analyses of WMH on processing speed, global function and global cognition, using temporal meta-ROIs.
3. Fig S3. Alternative mediation models tested.
4. Table S1. Inclusion and exclusion criteria for MITNEC-C6.
5. Table S2. Demographics in the MITNEC-C6 and ADNI cohorts.
6. Table S3. Description of cognitive tests and functional activities.
7. Table S4. Description of MR imaging parameters.
8. Table S5. Lobar WMH volumes vs cognitions.

