## Supplementary materials for "Vascular burden and cognition: Mediating roles of neurodegeneration and amyloid-PET"

Figures:

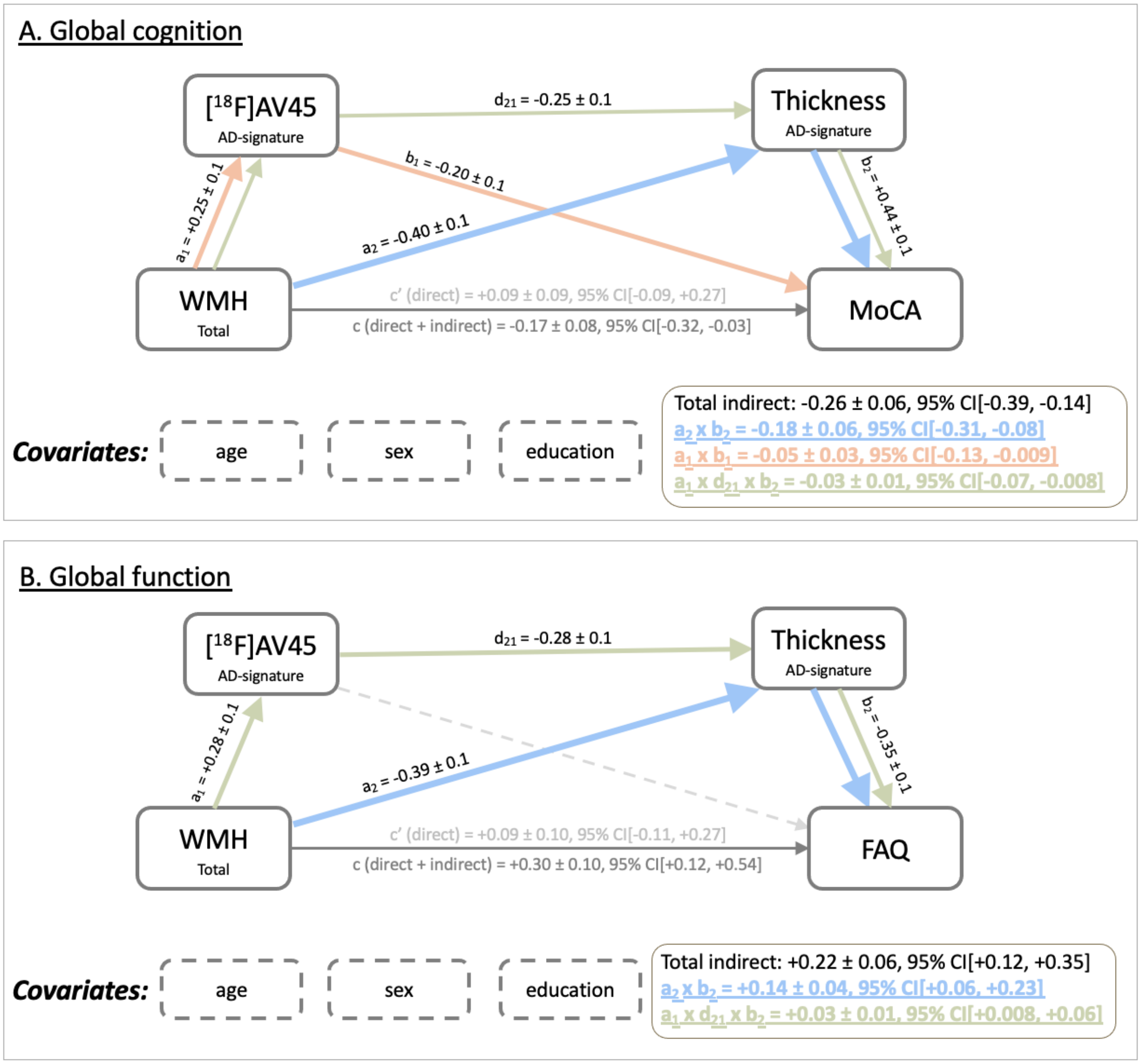

**Figure S1. Mediation analyses of Aß and atrophy on the WMH-cognition relationship.** Mediation models showing significant paths mediating the association of total WMH volumes with global cognition (MoCA; Panel A) and global function (FAQ; Panel B). Thick lines are part of a significant pathway, whereas dashed lines represent non-significant pathways. All mediators used an AD-signature meta-ROI. Values are indicated as mean ± SE and 95% CI are bootstrapped with 5,000 replications. Path c represents the total (direct + indirect) effect adjusted only for covariates, whereas c' represents the direct effect adjusted for covariates and indirect effects.

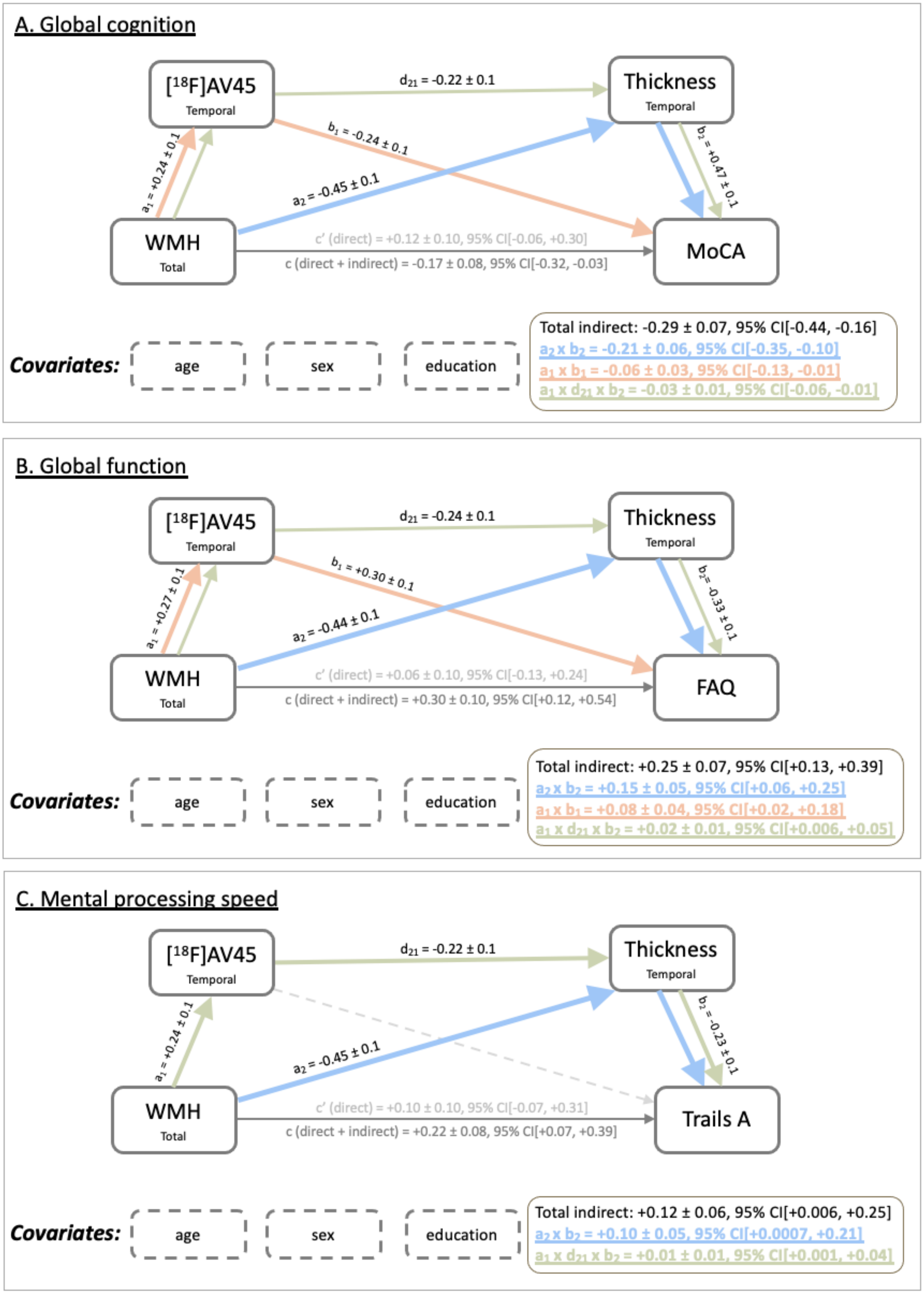

**Figure S2. Mediation analyses of Aß and atrophy on the WMH-cognition relationship.** Mediation models showing significant paths mediating the association of total WMH volumes with global cognition (MoCA; Panel A), global function (FAQ; Panel B), and mental processing speed (TrailsA, Pancel C). Thick lines are part of a significant pathway, whereas dashed lines represent non-significant pathways. All mediators used a temporal meta-ROI. Values are indicated as mean ± SE and 95% CI are bootstrapped with 5,000 replications. Path c represents the total (direct + indirect) effect adjusted only for covariates, whereas c' represents the direct effect adjusted for covariates and indirect effects.

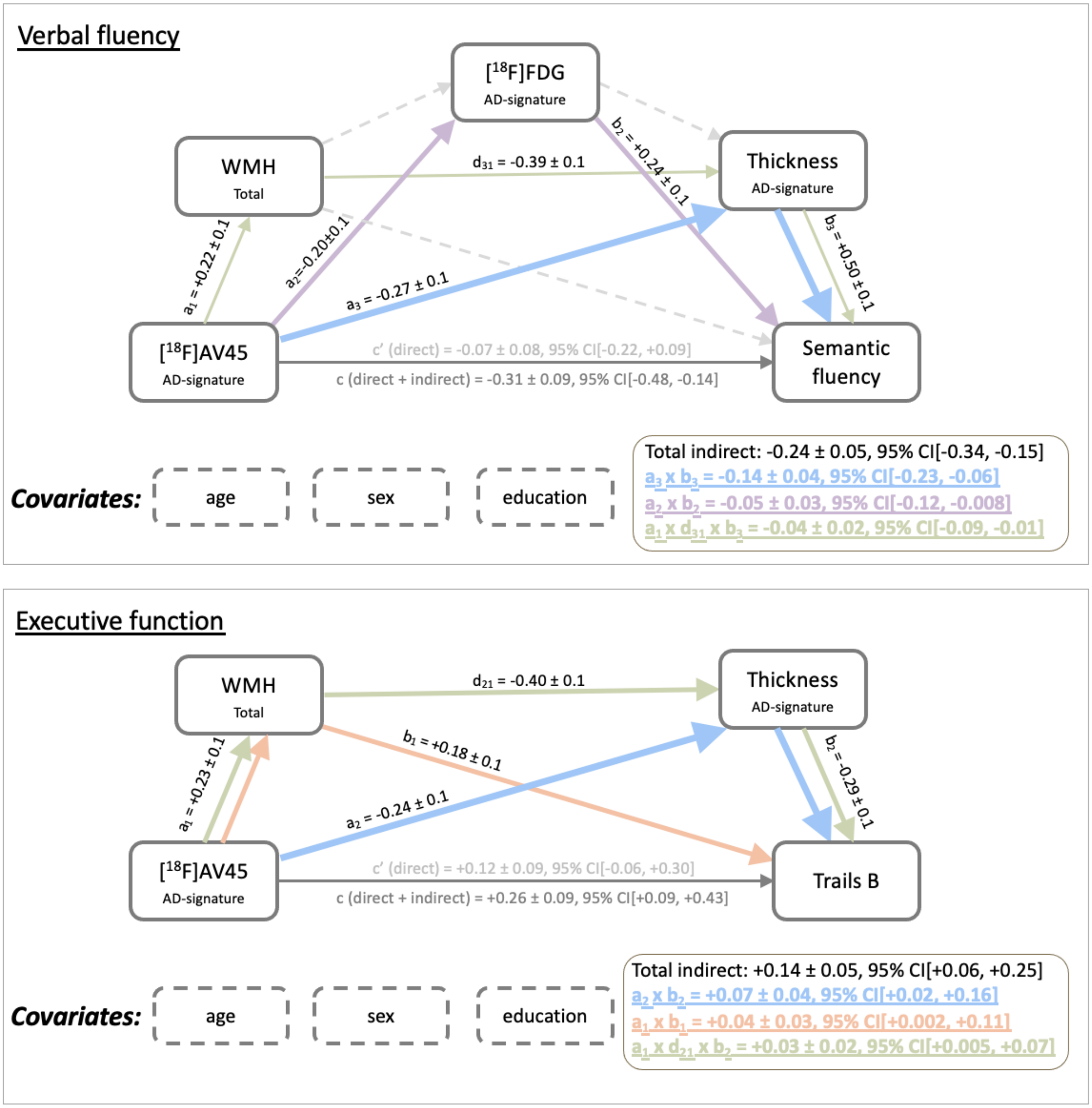

**Figure S3. Post-hoc mediation analyses using Aß as predictor and WMH as mediator.** Thick lines are part of a significant pathway, whereas dashed lines represent non-significant pathways. All mediators used an AD-signature meta-ROI. Values are indicated as mean ± SE and 95% CI are bootstrapped with 5,000 replications. Path c represents the total (direct + indirect) effect adjusted only for covariates, whereas c' represents the direct effect adjusted for covariates and indirect effects.

### Tables:

**Table S1. Inclusion and Exclusion Criteria for MITNEC-C6.**

| Inclusion Criteria | Exclusion Criteria |
|---|---|
| 1. A. Early AD or amnestic, non-amnestic single or multi-domain MCI with confluent pvWMH; recruited from memory clinics or B. minor stroke (e.g., subcortical lacunar infarct ≤1.5cm) or a TIA with confluent pvWMH; recruited from stroke prevention clinics<br>2. Age ≥ 60<br>3. Written informed consent<br>4. More than 8 years of education<br>5. Expected survival greater than 2 years<br>6. Sufficiently fluent in French or English for cognitive testing<br>7. Mini-Mental State Exam score ≥ 20<br>8. pvWMH score on CT or MRI of ≥2 on the periventricular Fazekas scale* | 1. Cortical or non-lacunar infarct on imaging<br>2. Persisting hemiparesis after a motor stroke, leg strength <4/5 on the Medical Research Council (MRC) scale; significant cerebellar ataxia<br>3. Contraindications to 3T MRI<br>4. Major psychiatric disorder in the preceding 5 years<br>5. History of substance abuse in the past 2 years<br>6. Serious/chronic systemic or neurological illness (other than AD) such as Parkinson's disease, multi-infarct dementia, Huntington's disease, normal pressure hydrocephalus, brain tumor, progressive supranuclear palsy, seizure disorder, subdural hematoma, multiple sclerosis, or history of significant head trauma followed by persistent neurologic deficits or structural brain abnormalities<br>7. Pain or sleep disorder that could interfere with testing<br>8. Claustrophobia<br>9. Radiation therapy to the head or neck or in a research study involving radiation<br>10. Unable or unwilling to comply with protocol requirements or deemed by the investigator to be unfit for the study |

* Fazekas 2 can be included if bilateral posterior or anterior periventricular caps extending at least 10mm from the ventricle (i.e., halfway into surrounding white matter vs. extending out to most surrounding white matter for Fazekas 3)

**Table S2. Demographics.** All values are indicated as mean ± standard deviation except for sex and Aß-positivity ($^{*}P = 0.03$, $^{**}P < 0.001$). Abbreviations: BNT, Boston Naming Test; FAQ Functional Assessment Questionnaire; MMSE, mini-mental state examination; MoCA, Montreal Cognitive assessment; SUVR, standardized uptake value ratio; Trails, Trail Making Test; WMH, white matter hyperintensity volumes.

| **Variables** | **Cohort 1 (n = 60)** | **Cohort 2 (n = 60)** |
|---|---|---|
| **Recruitment** | ADNI-2 | MITNEC-C6[4] |
| **Age (years)** | 74.01 ± 5.47 | 76.85 ± 8.01$^{*}$ |
| **Sex male, n (%)** | 25 (42%) | 34 (57%) |
| **Education (years)** | 16.13 ± 2.73 | 14.25 ± 2.66$^{**}$ |
| **Pulse pressure (mmHg)** | 61.32 ± 14.35 (n=47) | 62.32 ± 16.02 (n=59) |
| **Hypertension[1], n (%)** | 28 (47%) | 35 (58%) |
| **BMI** | 27.65 | 27.36 |
| **Smoking history, n (%)** | 32 (53%) | 26 (43%) |
| **Composite [$^{18}$F]AV45 SUVR[2]** | 0.75 ± 0.35 | 1.08 ± 0.44$^{**}$ |
| **Aß-positive, n (%)** | 13 (22%) | 29 (48%) |
| **Composite [$^{18}$F]FDG SUVR[2]** | 2.30 ± 0.28 | 2.26 ± 0.36 |
| **Cortical thickness (global, mm)** | 2.33 ± 0.08 | 2.26 ± 0.10$^{**}$ |
| **Total WMH (cm$^3$)** | 10.61 ± 12.89 | 34.15 ± 18.80$^{**}$ |
| **Depression scale[3]** | 0.75 ± 1.10 (GDS) | 11.57 ± 10.04 (CES-D) |
| **Semantic fluency** | 20.95 ± 5.57 | 12.8 ± 5.93$^{**}$ |
| **Trails A (seconds)** | 36.30 ± 11.11 | 57.62 ± 32.81$^{**}$ |
| **Trails B (seconds)** | 95.78 ± 49.91 | 186.31 ± 84.79$^{**}$ (n=59) |
| **BNT** | 27.67 ± 2.14 | 23.72 ± 5.66$^{**}$ (n=54) |
| **FAQ** | 0.5 ± 1.23 | 6.36 ± 8.07$^{**}$ (n=50) |
| **MMSE** | 28.88 ± 1.35 | 27.08 ± 2.46$^{**}$ |
| **MoCA** | 25.62 ± 2.50 | 22.42 ± 4.39$^{**}$ |

[1] Hypertension was defined as systolic blood pressure ≥ 140 mmHg and/or diastolic blood pressure ≥ 90 mmHg.
[2] SUVR values were corrected for partial volume effects. Composite [$^{18}$F]AV45 and [$^{18}$F]FDG SUVR were based on Jack et al. 2008 *Brain* and Landau et al. 2011 *Neurobiol Aging*, respectively.
[3] For ADNI and MITNEC-C6, the Geriatric Depression Scale (GDS) and the Center for Epidemiologic Studies Depression scale (CES-D) were applied, respectively.
[4]MITNEC-C6 subjects were recruited from stroke-prevention (n=17) and dementia clinics (n=43)

**Table S3. Description of cognitive tests and functional activities.**

### 1. Trail Making tests (Trails-A and Trails-B) [1]

**DESCRIPTION**
This is a test of processing speed and executive function. Although both Parts A and B depend on visuomotor and perceptual-scanning skills, Part B also requires considerable cognitive flexibility in shifting from number to letter sets under time pressure.
**PART A** consists of 25 circles numbered 1 through 25 distributed over a white sheet. The participant is instructed to connect the circles with a drawn line as quickly as possible in ascending numerical order.
**PART B** also consists of 25 circles, but these circles contain either numbers (1 through 13) or letters (A through L). The participant must connect the circles while alternating between numbers and letters in ascending order (e.g., A to 1; 1 to B; B to 2; 2 to C).
The participant's performance is judged in terms of the time, in seconds, required to complete each Trail.

### 2. Boston Naming Test (BNT) [2]

**DESCRIPTION**
This test is a measure of the ability to orally label (name) thirty line drawings of objects. The objects are presented in order of frequency, from most frequent (i.e., bed) to least frequent (i.e., protractor). This test is sensitive to aphasia and also to object recognition deficits.

### 3. Semantic fluency [3]

**DESCRIPTION**
This is a short test of category fluency. The participant is given one minute to produce as many unique words as possible within a semantic category: 'name any member of the animal kingdom'. The participant's score in each task is the number of unique correct words.

### 4. Montreal cognitive assessment (MoCA) [4]

**DESCRIPTION**
The MoCA assesses different cognitive subdomains, including short term memory, visuospatial abilities, executive functions, attention, concentration and working memory, language, and orientation to time and place. The total test score corresponds to 30 points. The MoCA is often used as a screening test to detect MCI or AD.

### 5. Functional assessment questionnaire (FAQ) [5]

**DESCRIPTION**
This test measures instrumental activities of daily living. It measures the representation of the participant's level of ability to perform certain daily activities over the preceding four weeks. The FAQ is often used in clinical settings as a screening test to detect MCI or AD or to discriminate between both.

**Table S4. Description of MR imaging parameters.**

| Study | MITNEC – C6 | | |
|---|---|---|---|
| Sequence | 3DT1 | | |
| ***Protocol*** | | | |
| Vendor | GE | Philips | Siemens |
| Field Strength | 3T | 3T | 3T |
| Model | Discovery | Achieva | Skyra/Trio/Prisma |
| Version | 22 | 3.2.3 | |
| Sequence Name | 3D FAST SPGR | 3D TFE | 3D MP-RAGE |
| Imaging Options | IrP- Asset | Fast (Sense) | iPat |
| ***Pulse Timing*** | | | |
| TE (ms) | Min full | Min (3.3) | 2.98 |
| TR (ms) | Min | Min (7.3) | 2300 |
| Flip Angle (°) | 11 | 9 | 9 |
| TI (ms) | 400 | 945 | 900 |
| ***Scan Range*** | | | |
| FOV (in-plane) (mm) | 256 x 256 | 256 x 248 | 256 x 256 |
| Slice Thickness (mm) | 1 | 1 | 1 |
| Gap Between Slices (mm) | 0 | 0 | 0 |
| No. Slices | 176 | 176 | 176 |
| ***Acquisition*** | | | |
| Orientation | Sagittal | Sagittal | Sagittal |
| Matrix Size | 256 x 256 | 256 x 248 | 256 x 256 |
| Voxel Size [L/R x A/P x I/S] | 1 x 1 x 1 | 1 x 1 x 1 | 1 x 1 x 1 |
| NEX | 1 | 1 | 1 |
| Acceleration Factor (Parallel factor) | 2 | 2 | 2 |
| ***Other*** | | | |
| Fat Suppression | None | None | None |
| Bandwidth | 31.25 (kHz) | 228 (Hz/px) | 240 (Hz/px) |
| Echo Train Length | - | - | - |
| ***Coil Type*** | | | |
| Head | X | X | X |
| Channel | 8-12 (HNS) | 8 | 12(Trio) 20 (Prisma) |

| Sequence | 2D FLAIR | | |
|---|---|---|---|
| ***Protocol*** | | | |
| Sequence Name | 2D T2FLAIR | 2D IR TSE | 2D IR TDF |
| Imaging Options | EDR, IR | Fast (Sense) | iPat |
| ***Pulse Timing*** | | | |
| TE (ms) | 140 | 125 | 120 |
| TR (ms) | 9000 | 9000 | 9000 |
| Flip Angle (°) | 125 | 90 (150 refocus) | 165 |

| | | | |
|---|---|---|---|
| TI (ms) | 2250 | 2500 | 2500 |
| ***Scan Range*** | | | |
| FOV (in-plane) (mm) | 240 x 240 | 240 x 240 | 240 x 240 |
| Slice Thickness (mm) | 3 | 3 | 3 |
| Gap Between Slices (mm) | 0 | 0 | 0 |
| No. Slices | 48 | 48 | 48 |
| ***Acquisition*** | | | |
| Orientation | Oblique Axial | Oblique Axial | Oblique Axial |
| Matrix Size | 256 x 256 | 256 x 242 | 256 x 256 |
| Voxel Size [L/R x A/P x I/S] | 0.94 x 0.94 x 3 | 0.94 x 0.99 x 3 | 0.94 x 0.94 x 3 |
| NEX | 1 | 1 | 1 |
| Acceleration Factor (Parallel factor*) | No Asset | 2 (SENSE) | 2 |
| ***Other*** | | | |
| Fat Suppression | None | None | None |
| Bandwidth | 25 (kHz) | 242 (Hz/px) | 220 (Hz/px) |
| Echo Train Length | | 19 | 19 |
| ***Coil Type*** | | | |
| Head | X | X | X |
| Channel | 8-12 (HNS) | 8 | 12(Trio) 20 (Prisma) |

ADNI-2: http://adni.loni.usc.edu/wp-content/uploads/2010/05/ADNI2_GE_3T_22.0_T2.pdf

**Table S5. Lobar WMH volumes vs. cognitions.** All values are indicated as mean ± SE and ranked in order of effect size. P-values are bootstrapped with 5,000 replications and adjusted for age, sex, and education ($^{*}$P<0.05, $^{**}$P<0.01, $^{***}$P<0.001). Non-significant associations between WMH and cognitive metrics across all subjects were omitted from the table. Abbreviations: *A*ß+/- Aß-positive (n=42) / Aß-negative (n=78); BNT, Boston Naming Test; FAQ, functional assessment questionnaire; MoCA, Montreal cognitive assessment; n.s., non-significant; WMH, white matter hyperintensity volumes.

| WMH | Cognition | All | *A*ß+ group | *A*ß- group |
|---|---|---|---|---|
| ***Total*** | Semantic fluency | -0.35 ± 0.09$^{***}$ | -0.60 ± 0.19$^{**}$ | -0.25 ± 0.10$^{*}$ |
| | Executive function (trails B) | +0.34 ± 0.09$^{***}$ | +0.48 ± 0.21$^{*}$ | +0.28 ± 0.10$^{**}$ |
| | Global function (FAQ) | +0.30 ± 0.10$^{**}$ | +0.72 ± 0.28$^{*}$ | n.s. |
| | Processing speed (trails A) | +0.22 ± 0.08$^{*}$ | n.s. | n.s. |
| | Global cognition (MoCA) | -0.17 ± 0.08$^{*}$ | n.s. | n.s. |
| ***Frontal*** | Semantic fluency | -0.35 ± 0.08$^{***}$ | n.s. | -0.38 ± 0.09$^{***}$ |
| | Executive function (trails B) | +0.35 ± 0.09$^{***}$ | n.s. | +0.43 ± 0.10$^{***}$ |
| | Global function (FAQ) | +0.32 ± 0.12$^{**}$ | n.s. | n.s. |
| | Processing speed (trails A) | +0.28 ± 0.10$^{**}$ | n.s. | +0.35 ± 0.15$^{*}$ |
| ***Parietal*** | Executive function (trails B) | +0.29 ± 0.09$^{**}$ | n.s. | +0.28 ± 0.12$^{*}$ |
| | Semantic fluency | -0.27 ± 0.09$^{**}$ | -0.32 ± 0.15$^{*}$ | n.s. |
| ***Cingulate*** | Executive function (trails B) | +0.36 ± 0.09$^{***}$ | +0.40 ± 0.19$^{*}$ | +0.31 ± 0.11$^{**}$ |
| | Semantic fluency | -0.31 ± 0.09$^{**}$ | n.s. | -0.22 ± 0.11$^{*}$ |
| | Global cognition (MoCA) | -0.22 ± 0.08$^{**}$ | -0.35 ± 0.17$^{*}$ | n.s. |
| | Language (BNT) | -0.22 ± 0.08$^{*}$ | -0.45 ± 0.20$^{*}$ | n.s |
| | Global function (FAQ) | +0.18 ± 0.09$^{*}$ | n.s. | n.s. |
| | Processing speed (trails A) | +0.17 ± 0.08$^{*}$ | n.s. | n.s. |
| ***Temporal*** | Executive function (trails B) | +0.30 ± 0.11$^{**}$ | +0.57 ± 0.16$^{***}$ | +0.22 ± 0.12$^{*}$ |
| | Semantic fluency | -0.23 ± 0.11$^{*}$ | -0.39 ± 0.20$^{*}$ | n.s. |
| ***Insula*** | Semantic fluency | -0.33 ± 0.08$^{***}$ | n.s. | -0.32 ± 0.09$^{***}$ |
| | Executive function (trails B) | +0.20 ± 0.09$^{*}$ | n.s. | +0.23 ± 0.11$^{*}$ |
